# Chronotype and mortality - a 37-year follow-up study in Finnish adults

**DOI:** 10.1101/2022.04.02.22273342

**Authors:** Christer Hublin, Jaakko Kaprio

## Abstract

**Background:** The only study on chronotype and mortality suggested small increases of all-cause and cardiovascular mortality in a 6.5-year follow-up. Our aim was to constructively replicate findings from it in a longer follow-up.

**Methods:** A questionnaire was administered to the population-based Finnish Twin Cohort in 1981 (response rate 84%, age 24–101 years). The study population included 23,854 participants who replied to the question: “Try to assess to what extent you are a morning person or an evening person”, with four response alternatives (clearly a morning person, to some extent a morning person, to some extent an evening person, and clearly an evening person). Vital status and cause of death data were provided by nationwide registers up to the end of 2018. Hazard ratios and their 95% confidence intervals for mortality were computed; there were 8728 deaths over the follow-up period. Adjustments were made for education, alcohol, smoking, body mass index, and sleep length.

**Results:** The model adjusted for all covariates showed a 9% increase in risk of all-cause mortality for the evening-type group (1.09, 1.01–1.18), with attenuation mainly due to smoking and alcohol, as they had larger consumption than those with morningness chronotypes. We observed no increase in cardiovascular mortality by chronotype. No increase in mortality was seen among the non-smokers who were at most light drinkers.

**Conclusions:** Our results from this 37-year follow-up study suggest that there is little or no independent contribution of chronotype to all-cause and cardiovascular mortality.

**Key messages:**

- There is little or no independent contribution of chronotype to mortality.
- The increased risk of mortality associated with eveningness appears to be mainly mediated by a larger consumption of tobacco and alcohol than in those with a morningness chronotype.
- It is important to be aware of the increased health risks of evening-type persons.

## Introduction

Good sleep is essential for health. Chronically deficient sleep, manifested as a reduced amount or quality of sleep or mistiming of sleep, is associated with many negative health outcomes^1^. Genetic factors influence many aspects of sleep, including sleep quality^2^, sleep length^3^, and chronotype^4^ (also called diurnal type). Chronotype is the phenotypic expression of an individual’s innate circadian rhythm, that is the part of the day between morning and evening that is preferred for daily activities^5^. The later chronotype (evening type) is associated with an evening preference for activities and a later timing of sleep.

The later chronotype has been associated with increased morbidity, including impaired cardiometabolic health^6-8^ and increased risk of psychiatric symptoms^9,10^. Additionally, the tendency towards eveningness is associated with unhealthy dietary habits^11^ and overweight^12^, as well as increased frequency of smoking^13^ and alcohol use^14^. Additionally, the evening chronotype is associated with poor work ability and disability pensions at midlife^15^. Given the associations of chronotype with lifestyle factors that are known to increase the risk of premature morbidity and mortality, the independent contribution of chronotype to mortality is of relevance when providing public health recommendations related to sleep and chronotype.

Recently, a large cohort study based on the UK Biobank (UKBB) suggested a small increased risk of all-cause mortality (2%) and cardiovascular mortality (4%)^16^. They followed 433,268 men and women aged 38–73 years at baseline, with 10,534 deaths occurring during an average follow-up of 6.5 years. Multiple covariates were considered in the analyses.

Our aim was to constructively replicate findings from the UK Biobank study in a cohort that also included younger adults at baseline and with a much longer follow-up. To study the association between chronotype and all-cause mortality and mortality from selected disease groups, we examined mortality between 1981 and 2018 in relation to chronotype assessed at baseline in 1981 in a large population-based cohort of twin individuals aged 24 years and over from Finland.

## Methods

### Population sample

The Older Finnish Twin Cohort is a longitudinal study of Finnish twin pairs of the same sex born before 1958, who were alive in 1975^17^. As these pairs were selected from the Population Information System, Digital and Population Data Services Agency of Finland (dvv.fi) in 1974, the cohort is population-based, and its overall mortality and cancer incidence does not differ from that of the general population^18,19^. The 1981 questionnaire yielded a response rate of 84% (N = 24,945 individuals). The questionnaire included about 100 questions on demographic, social, health/illness and lifestyle variables.

The study has been approved by the ethical committee of the Department of Public Health, University of Helsinki. Informed consent was obtained from all respondents. The current record linkage to cause-of-death data was approved by Statistics Finland (permit Dnro TK-53-1608-19) and by the Research Ethics Board, Finnish Institute for Health and Welfare (THL 220/6.02.04/2021).

### Questionnaire and register data

The chronotype was asked by a question according to the Diurnal Type Scale^20^: “Try to assess to what extent you are a morning person or an evening person”. There were four reply alternatives: “I am clearly a morning person” (morning type); “I am to some extent a morning person” (somewhat morning type); “I am to some extent an evening person” (somewhat evening type); “I am clearly an evening person” (evening type)^4^. In our mortality analyses, we considered the morning persons as the reference category.

Additionally, we asked attained educational level (as a proxy for socioeconomic status), alcohol consumption, smoking status, weight and height to compute body mass index (BMI, kg/m^2^), and sleep length (for categories see Table 1)^21^. These are included as potential confounders and known predictors of mortality.

**Table 1.** Baseline characteristics in 1981 questionnaire of the Finnish Twin Cohort by chronotype.

|  | Morning type | Some morning type | Some evening type | Evening type | Total |
| --- | --- | --- | --- | --- | --- |
|  | N=6,769 | N=6,354 | N=7,591 | N=2,262 | N=22,976 |
| Sex |  |  |  |  |  |
| Women | 3,653 (54.0%) | 3,123 (49.2%) | 3,956 (52.1%) | 1,162 (51.4%) | 11,894 (51.8%) |
| Age [mean (SD)] in years | 45.5 (14.3) | 40.3 (12.9) | 37.1 (11.7) | 38.9 (13.2) | 40.6 (13.4) |
| Education (years) |  |  |  |  |  |
| Under 6 (Less than primary school) | 300 (4.4%) | 133 (2.1%) | 134 (1.8%) | 57 (2.5%) | 624 (2.7%) |
| 6 (Primary school) | 3,384 (50.0%) | 2,545 (40.1%) | 2,727 (35.9%) | 739 (32.7%) | 9,395 (40.9%) |
| 7-8 (Primary school and some vocational training) | 1,619 (23.9%) | 1,790 (28.2%) | 2,081 (27.4%) | 535 (23.7%) | 6,025 (26.2%) |
| 9 (Middle school) | 298 (4.4%) | 308 (4.8%) | 444 (5.8%) | 137 (6.1%) | 1,187 (5.2%) |
| 10-11 (Middle school and some vocational training) | 477 (7.0%) | 608 (9.6%) | 823 (10.8%) | 270 (11.9%) | 2,178 (9.5%) |
| 12 (High school) | 47 (0.7%) | 71 (1.1%) | 121 (1.6%) | 54 (2.4%) | 293 (1.3%) |
| 13-14 (High school and some vocational training) | 248 (3.7%) | 398 (6.3%) | 574 (7.6%) | 208 (9.2%) | 1,428 (6.2%) |
| 15 or more (University) | 252 (3.7%) | 343 (5.4%) | 480 (6.3%) | 204 (9.0%) | 1,279 (5.6%) |
| Other | 144 (2.1%) | 158 (2.5%) | 207 (2.7%) | 58 (2.6%) | 567 (2.5%) |
| Alcohol consumption (grams/day) |  |  |  |  |  |
| None | 2,265 (33.5%) | 1,529 (24.1%) | 1,535 (20.2%) | 497 (22.0%) | 5,826 (25.4%) |
| 1-10 | 3,345 (49.4%) | 3,668 (57.7%) | 4,455 (58.7%) | 1,170 (51.7%) | 12,638 (55.0%) |
| 11-20 | 571 (8.4%) | 584 (9.2%) | 826 (10.9%) | 262 (11.6%) | 2,243 (9.8%) |
| 21-30 | 221 (3.3%) | 254 (4.0%) | 309 (4.1%) | 115 (5.1%) | 899 (3.9%) |
| 31-40 | 146 (2.2%) | 137 (2.2%) | 200 (2.6%) | 81 (3.6%) | 564 (2.5%) |
| 41 or more | 221 (3.3%) | 182 (2.9%) | 266 (3.5%) | 137 (6.1%) | 806 (3.5%) |
| Smoking status (Cigarettes Per Day) |  |  |  |  |  |
| Never | 3,494 (51.6%) | 3,107 (48.9%) | 3,435 (45.3%) | 851 (37.6%) | 10,887 (47.4%) |
| Occasional | 201 (3.0%) | 217 (3.4%) | 254 (3.3%) | 77 (3.4%) | 749 (3.3%) |
| Former | 1,230 (18.2%) | 1,090 (17.2%) | 1,182 (15.6%) | 344 (15.2%) | 3,846 (16.7%) |
| Light (1-9 CPD) | 498 (7.4%) | 568 (8.9%) | 812 (10.7%) | 209 (9.2%) | 2,087 (9.1%) |
| Medium (10-19 CPD) | 841 (12.4%) | 885 (13.9%) | 1,288 (17.0%) | 443 (19.6%) | 3,457 (15.0%) |
| Heavy (20 or more CPD) | 505 (7.5%) | 487 (7.7%) | 620 (8.2%) | 338 (14.9%) | 1,950 (8.5%) |
| BMI [mean (SD)] | 24.4 (3.5) | 23.8 (3.4) | 23.4 (3.3) | 23.5 (3.6) | 23.8 (3.4) |
| Sleep length (hours) |  |  |  |  |  |
| 6 or less | 597 (8.8%) | 357 (5.6%) | 458 (6.0%) | 268 (11.8%) | 1,680 (7.3%) |
| 6½ | 478 (7.1%) | 437 (6.9%) | 538 (7.1%) | 240 (10.6%) | 1,693 (7.4%) |
| 7 | 1,364 (20.2%) | 1,402 (22.1%) | 1,783 (23.5%) | 549 (24.3%) | 5,098 (22.2%) |
| 7½ | 906 (13.4%) | 1,189 (18.7%) | 1,327 (17.5%) | 287 (12.7%) | 3,709 (16.1%) |
| 8 | 2,084 (30.8%) | 1,907 (30.0%) | 2,131 (28.1%) | 520 (23.0%) | 6,642 (28.9%) |
| 8½ | 545 (8.1%) | 536 (8.4%) | 645 (8.5%) | 135 (6.0%) | 1,861 (8.1%) |
| 9 | 556 (8.2%) | 395 (6.2%) | 500 (6.6%) | 155 (6.9%) | 1,606 (7.0%) |
| 9½ | 111 (1.6%) | 70 (1.1%) | 96 (1.3%) | 40 (1.8%) | 317 (1.4%) |
| 10 or more | 128 (1.9%) | 61 (1.0%) | 113 (1.5%) | 68 (3.0%) | 370 (1.6%) |

Vital status (alive in Finland on December 31, 2018, date of death or date of migration from Finland) as well as date of birth and sex provided at the formation of the cohort) were obtained from the Population Information System (dvv.fi), Finland. Cause of death data was provided by Statistics Finland. The follow-up for all-cause mortality and mortality from selected disease groups (ischaemic heart diseases, other heart diseases, cerebrovascular diseases, other diseases of the circulatory system, and dementia and Alzheimer’s disease deaths, see Table S3; alcohol deaths, see Table S4; lung cancer deaths, see Table S5) was from the exact date of response (date when questionnaire was returned) to December 31, 2018.

### Statistical analyses

Of the 24,945 individuals that replied to the 1981 survey, 4% did not reply to the chronotype item, leaving 23,854 persons to analyse. Of the respondents to the item on chronotype, 2.6% (626) were not included in the mortality analysis as they were living abroad or their address data was missing and hence it was unclear if they were resident in Finland at the start of follow-up. As only 1.1% of observations with data on chronotype and mortality had missingness for covariates (education, alcohol, smoking, BMI, and sleep length), we used a complete case analysis and no imputation was carried out. Over the follow-up period 8728 deaths occurred, with 14,248 observations censored (i.e., alive at end of follow-up or migration abroad). Follow-up time consisted of 551,124 person-years. Our focus was on the effect sizes and not statistical significance testing.

## Results

Of the respondents, 29.5% were morning types, 27.7% somewhat morning types, 33.0% somewhat evening types, and 9.9% evening types. Baseline characteristics with distributions of covariates are given in Table 1.

Table 2 shows risks of mortality by chronotype during the follow-up from 1981 to 2018. The age- and sex-adjusted model showed a 9% increased risk in the somewhat-evening-type group (hazard ratio (HR) 1.09, 95%confidence interval (CI) 1.03–1.15) and 21% increased risk in the evening-type group (1.21, 1.12–1.31). The model adjusted for all covariates shows an attenuated 9% increased risk for the evening-type group (1.09, 1.01– 1.18). The point estimates were similar, but with wider confidence intervals, when the sexes were analysed separately. For detailed statistics of the mortality associated with each covariate, see Table S2. Figure 1 shows the survival curves separately for men and women by chronotype.

**Table 2.** Risk of mortality by chronotype during the follow-up from 1981 to 2018. Four models with adjustment for age and sex only, adjustment for all covariates, adjusted for all covariates among men, and adjusted for all covariates among women. Risk given as hazard ratios with reference to morning type, 95% confidence intervals and p-values.

| Chronotype | Age and sex adjusted | Adjusted for all covariates | Adjusted for all covariates: Men | Adjusted for all covariates: Women |
| --- | --- | --- | --- | --- |
|  | HR<br>95% CI<br>p-value | HR<br>95% CI<br>p-value | HR<br>95% CI<br>p-value | HR<br>95% CI<br>p-value |
| Morning type (Reference) | 1.00<br>1.00, 1.00<br>- | 1.00<br>1.00, 1.00<br>- | 1.00<br>1.00, 1.00<br>- | 1.00<br>1.00, 1.00<br>- |
| Some morning type | 0.99<br>0.94, 1.04<br>0.693 | 1.00<br>0.95, 1.06<br>0.969 | 1.00<br>0.93, 1.07<br>0.902 | 1.00<br>0.92, 1.08<br>0.992 |
| Some evening type | 1.09<br>1.03, 1.15<br>0.004 | 1.04<br>0.98, 1.11<br>0.168 | 1.05<br>0.96, 1.13<br>0.279 | 1.03<br>0.95, 1.12<br>0.441 |
| Evening type | 1.21<br>1.12, 1.31<br><0.001 | 1.09<br>1.01, 1.18<br>0.028 | 1.09<br>0.99, 1.21<br>0.091 | 1.05<br>0.93, 1.18<br>0.436 |
| Time at risk (person-years) | 551,124 | 551,124 | 261,696 | 289,427 |
| N of deaths | 8,728 | 8,728 | 4,759 | 3,969 |
| N of persons | 22,976 | 22,976 | 11,082 | 11,894 |

**Figure:**
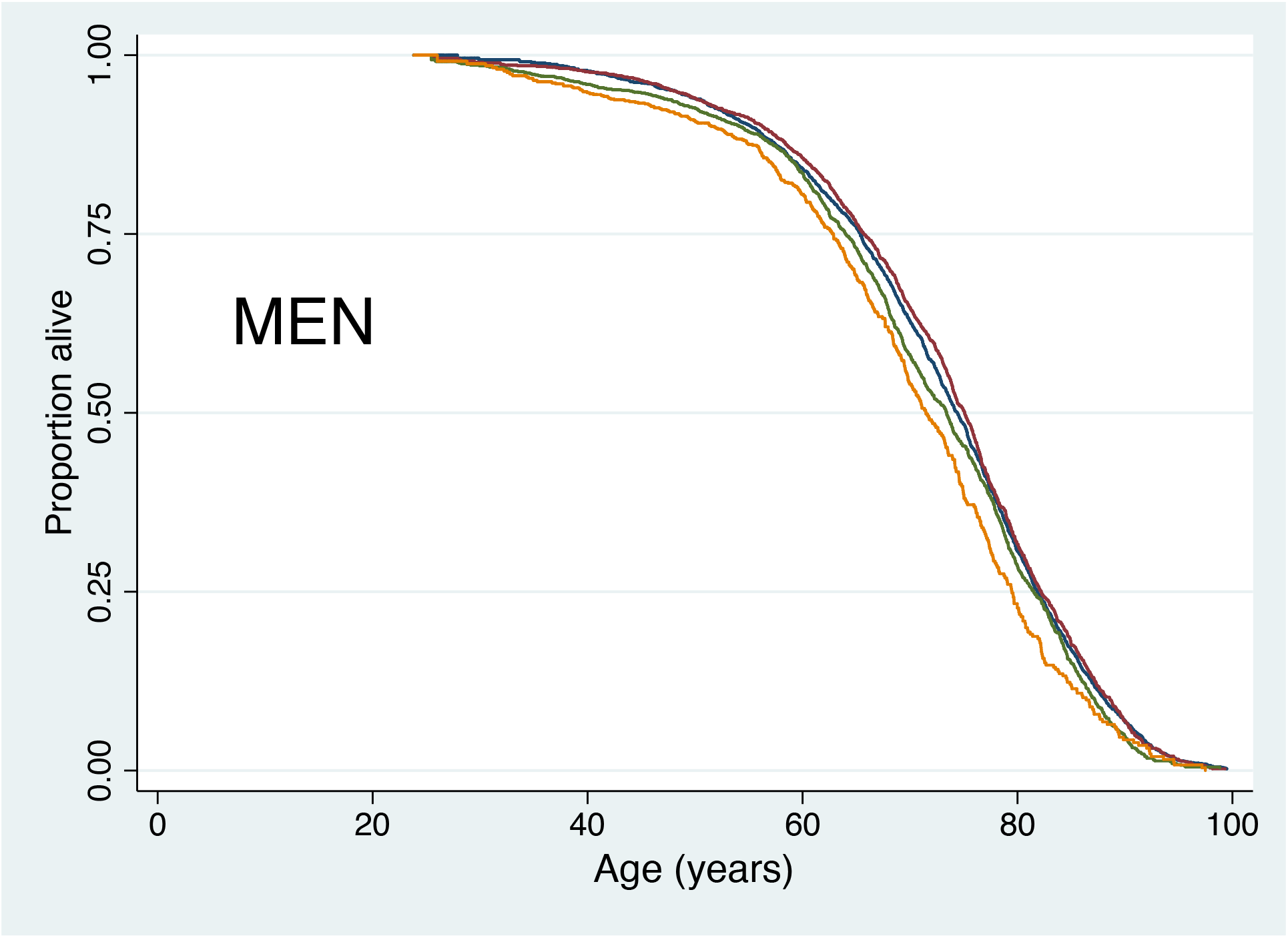
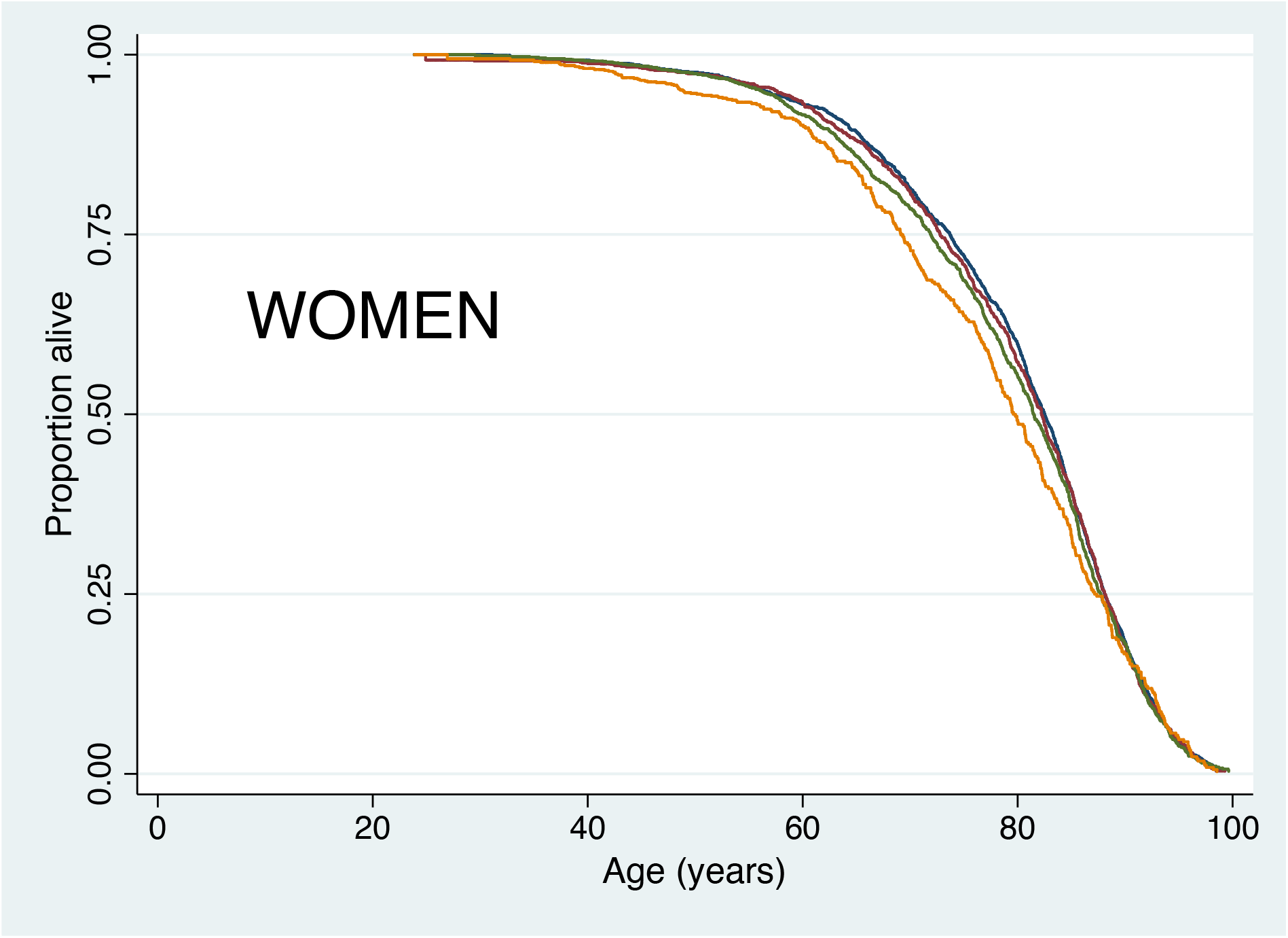
Survival curves for men and women in the Finnish Twin Cohort from 1981 to 2018 by chronotype (purple line is morning type, blue line somewhat morning type, green line is somewhat evening type and orange line is evening type)

The attenuation of risk in the evening-type group was mainly due to smoking and alcohol, which were also the strongest predictors of mortality after age and sex. As smoking and alcohol use are strongly correlated, we examined the risk by chronotype in groupings of alcohol and smoking. Table 3 gives age- and sex-adjusted mortality hazard ratios for chronotypes stratified by smoking status and alcohol consumption. In most subgroups, the HR estimates were close to one. Three elevated risk estimates (out of 36) were seen. The mortality of the somewhat-evening-type group was 47% increased among never smokers with moderate alcohol consumption (1.47, 1.07–2.03), though no effect was seen for evening-type in the same subgroup. In addition, the evening-type group showed higher HR estimates among former and occasional smokers (1.34, 1.07–1.68) and current smokers (1.22, 1.04–1.43), who were abstainers or light alcohol users.

**Table 3.** Age-sex adjusted mortality hazard ratios (and 95% confidence intervals and p-values) in 1981-2018 for chronotypes stratified by smoking status and alcohol consumption.

|  | Never smokers |  |  | Former and occasional smokers |  |  | Current smokers |  |  |
| --- | --- | --- | --- | --- | --- | --- | --- | --- | --- |
| Alcohol consumption (g/day) | 0-10 | 11-30 | 31 or more | 0-10 | 11-30 | 31 or more | 0-10 | 11-30 | 31 or more |
|  | HR<br>95%CI<br>p-value |  |  | HR<br>95%CI<br>p-value |  |  | HR<br>95%CI<br>p-value |  |  |
| Morning type | 1.00<br>1.00, 1.00<br>. | 1.00<br>1.00, 1.00<br>. | 1.00<br>1.00, 1.00<br>. | 1.00<br>1.00, 1.00<br>. | 1.00<br>1.00, 1.00<br>. | 1.00<br>1.00, 1.00<br>. | 1.00<br>1.00, 1.00<br>. | 1.00<br>1.00, 1.00<br>. | 1.00<br>1.00, 1.00<br>. |
| Some morning type | 1.01<br>0.93, 1.09<br>0.882 | 1.06<br>0.73, 1.53<br>0.763 | 1.13<br>0.63, 2.02<br>0.684 | 0.96<br>0.84, 1.09<br>0.487 | 1.05<br>0.79, 1.37<br>0.755 | 0.97<br>0.63, 1.51<br>0.904 | 1.01<br>0.90, 1.14<br>0.850 | 0.99<br>0.82, 1.18<br>0.897 | 0.92<br>0.74, 1.14<br>0.447 |
| Some evening type | 1.04<br>0.96, 1.13<br>0.359 | 1.47<br>1.07, 2.03<br>0.018 | 1.09<br>0.62, 1.92<br>0.753 | 0.99<br>0.86, 1.14<br>0.893 | 1.10<br>0.84, 1.44<br>0.493 | 1.09<br>0.75, 1.59<br>0.661 | 1.10<br>0.98, 1.25<br>0.109 | 0.94<br>0.79, 1.13<br>0.518 | 1.00<br>0.81, 1.23<br>0.964 |
| Evening type | 0.93<br>0.81, 1.05<br>0.248 | 0.99<br>0.59, 1.65<br>0.958 | 0.57<br>0.27, 1.17<br>0.1237 | 1.34<br>1.07, 1.68<br>0.011 | 0.93<br>0.64, 1.34<br>0.683 | 0.92<br>0.58, 1.46<br>0.717 | 1.22<br>1.04, 1.43<br>0.014 | 1.12<br>0.89, 1.40<br>0.347 | 1.14<br>0.90, 1.44<br>0.271 |
| Time at risk (person-years) | 24,3061 | 16,113 | 3,981 | 83,249 | 20,087 | 6,928 | 12,0464 | 40,441 | 20,203 |
| N of deaths | 3,621 | 201 | 74 | 1,303 | 338 | 165 | 1,769 | 778 | 568 |
| N of persons | 10,158 | 646 | 166 | 3,494 | 820 | 309 | 4,948 | 1,696 | 899 |

An age- and sex-adjusted analysis of those with missing response to chronotype produced similar HRs for mortality as for evening types (Table S1). Within-pair analysis showed a high degree of similarity between the co-twins (Table S1), and there was no effect on mortality of chronotype when comparing a twin with their co-twin. We observed no differences between the chronotypes in mortality from selected disease groups, including cardiovascular diseases and dementias (Table S3). Mortality from alcohol-related diseases and accidental poisoning by alcohol was greatly increased in the evening-type group (1.92, 1.33–2.76). Once alcohol use was included in the model, the hazard ratio for the evening-type group strongly attenuated and had an HR of 1.43 (0.97–2.10) in the model with all covariates (see Table S4). To assess the effect of smoking, we analysed mortality from malignant neoplasms of the larynx, trachea, bronchus and lung, which was increased in the evening-type group. After inclusion of smoking status, the evening type HR attenuated and was only slightly elevated in the model with all covariates (1.25, 0.90–1.74; see Table S5).

## Discussion

Our results showed a small (9%) increase in all-cause mortality during a 37-year follow-up in Finnish adults. We saw no increase for cardiovascular mortality, with hazard ratio point estimates that were below unity for ischaemic heart and cerebrovascular diseases in the evening-type group. Thus, our results differ somewhat from the recently published UK Biobank study suggesting a small (2%) increased risk of all-cause mortality and a larger 4% increase cardiovascular mortality in a 6.5-year follow-up^16^. However, alcohol was not analysed in that study, and only smoking status was adjusted for but not smoking quantity. As far as we know, these two are the only published epidemiological studies on chronotype and mortality, in addition to our paper on chronotype, shift work and prostate cancer^22^.

Given the attenuation of risk by smoking and alcohol – the strongest predictors of mortality after age and sex – the risk by chronotype was examined in strata of alcohol and smoking. In almost all of the 36 subgroups that were examined, the HR estimates were close to one; in particular among the non-smokers who were at most light drinkers, we saw no association of chronotype with mortality. The three subgroups with elevated risks are likely to be chance findings given that we could not see a consistent pattern over chronotypes. This indicates that there is little or no independent contribution of chronotype to mortality, and the increased risk of mortality associated with eveningness appears to be mainly mediated by a larger consumption of tobacco and alcohol than in those with morningness^23^. Greater nicotine dependence and greater smoking among evening types has been previously reported^13,24-26^. There is a reciprocal relationship between the reward system and circadian system, and the level of alcohol and substance use correlates with the preference to stay up later at night, i.e., eveningness^14,27^.

Further support for the important role of these two substances comes from examining causes of death that are strongly associated with smoking and alcohol use. Mortality from alcohol-related diseases and accidental poisoning by alcohol was increased among evening types (43%), but this effect was substantially attenuated when alcohol consumption was adjusted for. Expectedly, alcohol consumption linearly and strongly predicted deaths from alcohol-related diseases. Mortality from malignant neoplasms of the respiratory tract (used to assess the effect of smoking given that smoking accounts for a large majority of these cancers) was increased (25%) in evening types, however, this effect was again substantially attenuated when adjusted for smoking status and amount smoked. There were virtually no differences between the chronotypes in mortality from selected somatic disease groups, including cardiovascular diseases.

The studies have some similarities. Both used self-reports of the chronotype assessed with a single question with four options: in the present study Diurnal Type Scale^20^, and in the UKBB study a question similar to the last question of the Morningness-Eveningness Questionnaire^28^. Mortality information was obtained in both studies from comprehensive nationwide registers. The distribution of the chronotypes were similar, especially regarding the group of main interest – the evening types (9% in the UKBB study and 9.9% in our cohort).

The studies also have dissimilarities. The present study used the Older Finnish Twin Cohort as the study population, which is population-based, with overall mortality and cancer incidence not differing from that of the general population^17,19^. In the UKBB study, the cohort is population-based but generally healthier than the general UK population^29^, and the degree to which the findings are generalisable to the entire population, or to other countries, is not known^16,30^. The UKBB cohort is almost 20-fold larger in size compared to ours (433,268 vs. 23,854 adults). As the follow-up time is 5.7-fold longer in our study (6.5 vs. 37 years), this resulted in relatively similar numbers of deaths (10,534 vs. 8728). The covariates used in the models were largely similar in the studies, with one major exception. Alcohol was not included in the UKBB study, although it is a strong predictor of mortality^31,32^.

Our study has several strengths. Our cohort is truly population-based, making the results more generalisable to the entire population. The study population is relatively large, the follow-up time is long and the follow-up comprehensive. The long follow-up makes our results less vulnerable to reverse causation, that is subclinical morbidity changing the chronotype, other risk factors and at the same increasing mortality. We also had comprehensive data on lifestyle factors (including alcohol and smoking) known to be associated with chronotype^25^ and mortality as seen in the present analyses and reported earlier^33^. Our study also has some weaknesses. Chronotype is assessed using a single self-reported question, which is not similarly validated as the instruments most often used by circadian biologists, namely the Morningness-Eveningness Questionnaire^28^ and Munich Chronotype Questionnaire^34^. Both of these include several questions, making them less preferable to use in extensive questionnaires. Our question resembles the one used by Knutson and von Schantz^16^, and as they point out their question is practically identical to the final question of the Morningness-Eveningness Questionnaire with the highest correlation (p = 0.89) to the total score^35^.

In conclusion, our results suggest that there is little or no independent contribution of chronotype to mortality. The increased risk of mortality associated with eveningness would appear to be mainly mediated by a larger consumption of tobacco and alcohol than in those with morningness. It is important to be aware of the increased health risks of evening-type persons.

## Data Availability

The Finnish Twin Cohort (FTC) data is not publicly available due to the restrictions of informed consent. However, the FTC data is available through the Institute for Molecular Medicine Finland (FIMM) Data Access Committee (DAC) for authorised researchers who have IRB/ethics approval and an institutionally approved study plan. To ensure the protection of privacy and compliance with national data protection legislation, a data use/transfer agreement is needed, the content and specific clauses of which will depend on the nature of the requested data.

---

**Table S1.**
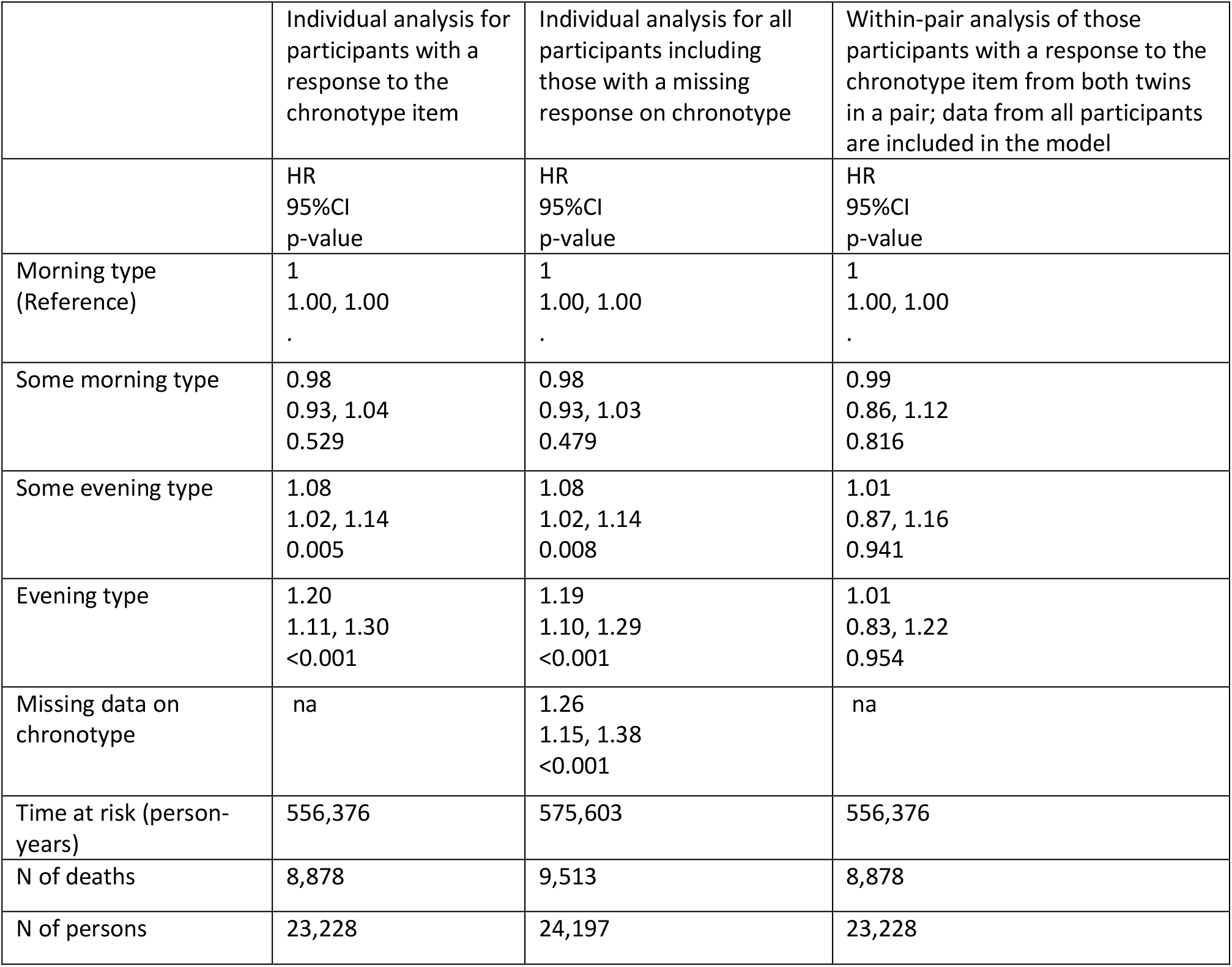
Age-sex adjusted hazard ratios, confidence intervals and p-values for three models of chronotype in 1981 and mortality 1981 to 2018.

**Table S2:** Hazard ratios, 95% confidence intervals and p-values for mortality 1981-2018 from multivariable models with all covariates in the same model. Models analyzed for all participants and for men and women separately.

|  | All | Men | Women |
| --- | --- | --- | --- |
|  | HR<br>95% CI<br>p-value | HR<br>95% CI<br>p-value | HR<br>95% CI<br>p-value |
| Morning type<br>(Reference) | 1.00<br>1.00, 1.00<br>. | 1.00<br>1.00, 1.00<br>. | 1.00<br>1.00, 1.00<br>. |
| Some<br>morning type | 1.00<br>0.95, 1.06<br>0.969 | 1.00<br>0.93, 1.07<br>0.902 | 1.00<br>0.92, 1.08<br>0.992 |
| Some<br>evening type | 1.04<br>0.98, 1.11<br>0.168 | 1.05<br>0.96, 1.13<br>0.279 | 1.03<br>0.95, 1.12<br>0.441 |
| Evening type | 1.09<br>1.01, 1.18<br>0.028 | 1.09<br>0.99, 1.21<br>0.091 | 1.05<br>0.93, 1.18<br>0.436 |
| Sex |  |  |  |
| Men<br>(Reference) | 1<br>1.00, 1.00<br>- | na | na |
| Women | 0.74<br>0.70, 0.78<br><0.001 | na | na |
| Education (years) |  |  |  |
| Under 6 (Less than primary<br>school) | 1<br>1.00, 1.00<br>. | 1<br>1.00, 1.00<br>. | 1<br>1.00, 1.00<br>. |
| 6 (Primary school) | 0.98<br>0.88, 1.08<br>0.651 | 0.96<br>0.83, 1.10<br>0.529 | 1.00<br>0.86, 1.17<br>0.980 |
| 7-8 (Primary school and some<br>vocational training) | 1.00<br>0.90, 1.12<br>0.974 | 1.01<br>0.87, 1.17<br>0.938 | 0.98<br>0.83, 1.16<br>0.815 |
| 9 (Middle school) | 0.95<br>0.82, 1.09<br>0.453 | 0.92<br>0.74, 1.16<br>0.489 | 0.94<br>0.76, 1.16<br>0.572 |
| 10-11 (Middle school and<br>some vocational training) | 0.93<br>0.81, 1.07<br>0.297 | 0.90<br>0.75, 1.08<br>0.247 | 0.97<br>0.79, 1.19<br>0.772 |
| 12 (High school) | 0.98<br>0.72, 1.34<br>0.920 | 1.02<br>0.67, 1.54<br>0.934 | 0.95<br>0.61, 1.50<br>0.834 |
| 13-14 (High school and some vocational training) | 0.96<br>0.80, 1.15<br>0.653 | 0.81<br>0.62, 1.06<br>0.120 | 1.10<br>0.86, 1.42<br>0.441 |
| 15 or more (University) | 0.81<br>0.69, 0.94<br>0.006 | 0.79<br>0.65, 0.96<br>0.015 | 0.84<br>0.67, 1.07<br>0.153 |
| Other | 0.94<br>0.78, 1.13<br>0.518 | 0.86<br>0.70, 1.07<br>0.179 | 1.23<br>0.83, 1.83<br>0.302 |
| Smoking (Cigarettes Per Day) |  |  |  |
| Never (Reference) | 1<br>1.00, 1.00<br>. | 1<br>1.00, 1.00<br>. | 1<br>1.00, 1.00<br>. |
| Occasional | 1.17<br>1.01, 1.36<br>0.036 | 1.10<br>0.91, 1.32<br>0.321 | 1.27<br>1.01, 1.60<br>0.045 |
| Former | 1.11<br>1.03, 1.18<br>0.005 | 1.03<br>0.95, 1.12<br>0.462 | 1.27<br>1.11, 1.44<br><0.001 |
| Light (1-9 CPD) | 1.55<br>1.40, 1.72<br><0.001 | 1.43<br>1.23, 1.65<br><0.001 | 1.69<br>1.46, 1.96<br><0.001 |
| Medium (10-19 CPD) | 1.97<br>1.83, 2.13<br><0.001 | 1.82<br>1.65, 2.00<br><0.001 | 2.11<br>1.86, 2.40<br><0.001 |
| Heavy (20 or more ) | 2.30<br>2.10, 2.51<br><0.001 | 2.05<br>1.86, 2.26<br><0.001 | 3.09<br>2.49, 3.85<br><0.001 |
| Alcohol use (grams/day) |  |  |  |
| Abstain | 1.00<br>0.95, 1.06<br>0.998 | 0.99<br>0.91, 1.09<br>0.899 | 1.01<br>0.94, 1.09<br>0.768 |
| 1-10 (Reference) | 1<br>1.00, 1.00<br>. | 1<br>1.00, 1.00<br>. | 1<br>1.00, 1.00<br>. |
| 11-20 | 1.24<br>1.14, 1.34<br><0.001 | 1.19<br>1.10, 1.29<br><0.001 | 1.53<br>1.24, 1.89<br><0.001 |
| 21-30 | 1.50<br>1.33, 1.69<br><0.001 | 1.47<br>1.30, 1.65<br><0.001 | 1.58<br>1.21, 2.06<br><0.001 |
| 31-40 | 1.75<br>1.51, 2.01<br><0.001 | 1.67<br>1.44, 1.93<br><0.001 | 2.16<br>1.38, 3.38<br><0.001 |
| 41 or more | 1.90<br>1.67, 2.17<br><0.001 | 1.87<br>1.64, 2.12<br><0.001 | 1.65<br>0.97, 2.81<br>0.067 |
| BMI | 1.00<br>0.99, 1.01<br>0.617 | 1.00<br>0.99, 1.01<br>0.469 | 0.99<br>0.98, 1.00<br>0.085 |
| Sleep length (hours) |  |  |  |
| 6 or less | 1.18<br>1.07, 1.30<br><0.001 | 1.22<br>1.06, 1.39<br>0.004 | 1.14<br>1.01, 1.30<br>0.037 |
| 6½ | 1.24<br>1.13, 1.36<br><0.001 | 1.19<br>1.05, 1.35<br>0.006 | 1.30<br>1.14, 1.49<br><0.001 |
| 7 | 1.10<br>1.02, 1.19<br>0.012 | 1.11<br>1.01, 1.23<br>0.034 | 1.09<br>0.97, 1.21<br>0.141 |
| 7½<br>(Reference) | 1<br>1.00, 1.00<br>. | 1<br>1.00, 1.00<br>. | 1<br>1.00, 1.00<br>. |
| 8 | 1.06<br>0.99, 1.14<br>0.119 | 1.01<br>0.92, 1.11<br>0.864 | 1.15<br>1.03, 1.27<br>0.009 |
| 8½ | 1.09<br>0.99, 1.21<br>0.094 | 1.11<br>0.97, 1.27<br>0.145 | 1.10<br>0.95, .27<br>0.202 |
| 9 | 1.18<br>1.07, 1.31<br>0.001 | 1.10<br>0.95, 1.27<br>0.189 | 1.30<br>1.13, 1.51<br><0.001 |
| 9½ | 1.07<br>0.89, 1.28<br>0.466 | 1.37<br>1.01, 1.85<br>0.040 | 0.94<br>0.74, 1.19<br>0.604 |
| 10 or more | 1.37<br>1.17, 1.61<br><0.001 | 1.33<br>1.06, 1.65<br>0.013 | 1.44<br>1.16, 1.80<br>0.001 |
| Healthy at baseline |  |  |  |
| No<br>(Reference) | 1<br>1.00, 1.00<br>. | 1<br>1.00, 1.00<br>. | 1<br>1.00, 1.00<br>. |
| Yes | 0.91<br>0.86, 0.95<br><0.001 | 0.87<br>0.82, 0.93<br><0.001 | 0.95<br>0.88, 1.02<br>0.122 |
| Time at risk (person-years) | 551,124 | 261,696 | 289,427 |
| N of deaths | 8,728 | 4,759 | 3,969 |
| N of persons | 22,976 | 11,082 | 11,894 |

**Table S3.**
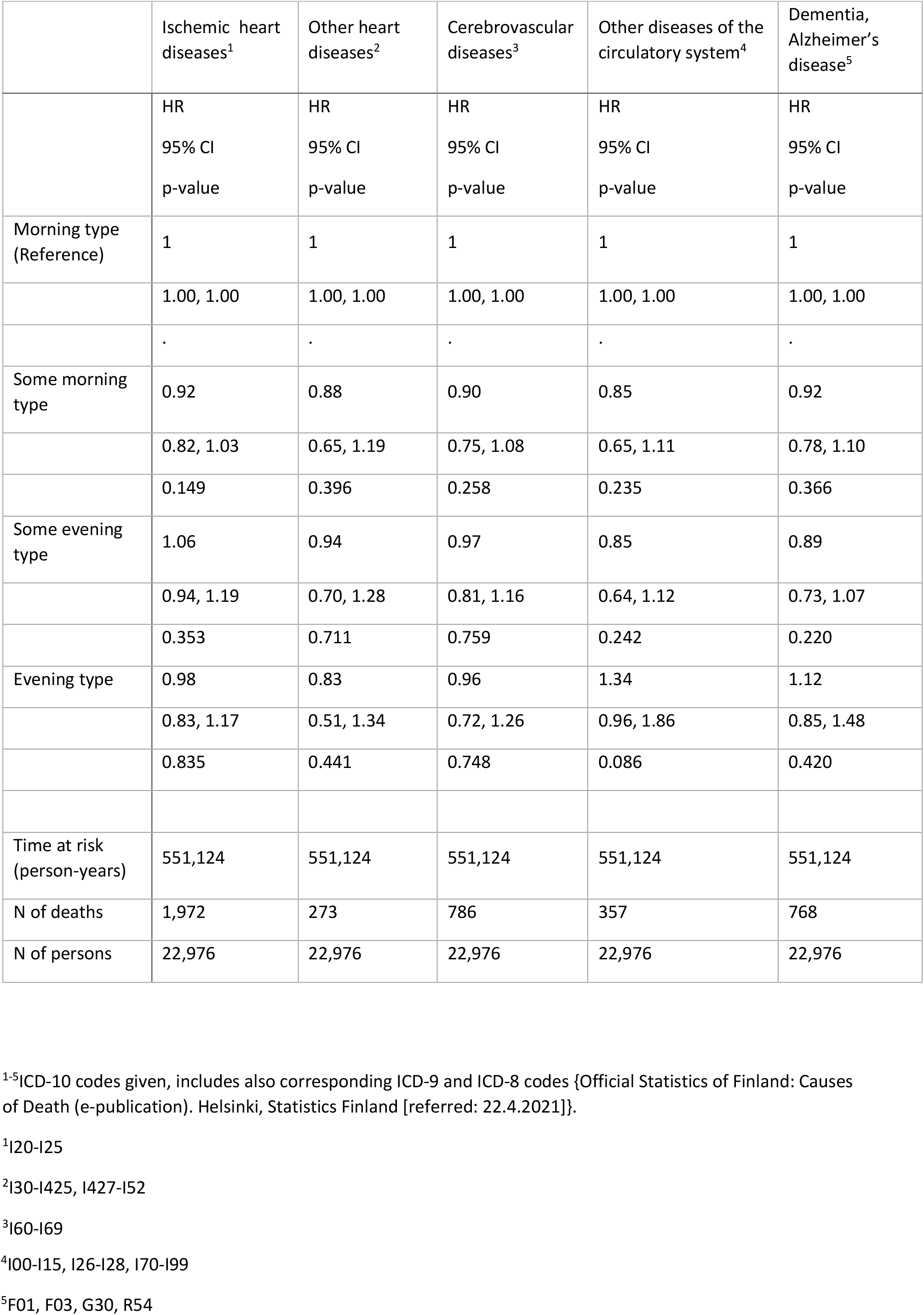
Hazard ratios, 95% confidence intervals and p-values from multivariable models for mortality from selected disease groups by chronotype in 1981-2018.

**Table S4.** Mortality from alcohol-related diseases and accidental poisoning by alcohol^1^: Hazard ratios, 95% confidence intervals and p-values from multivariable models for three models by chronotype and alcohol consumption in 1981-2018.

|  | Sex-age-adjusted model | Sex, age and alcohol consumption adjusted model | Model with all covariates, but only alcohol is shown |
| --- | --- | --- | --- |
|  | HR | HR | HR |
|  | 95% CI | 95% CI | 95% CI |
|  | p-value | p-value | p-value |
| Morning type (Reference) | 1 | 1 | 1 |
|  | 1.00, 1.00 | 1.00, 1.00 | 1.00, 1.00 |
|  | . | . | . |
| Some morning type | 1.16 | 1.15 | 1.12 |
|  | 0.85, 1.57 | 0.85, 1.55 | 0.83, 1.53 |
|  | 0.350 | 0.375 | 0.453 |
| Some evening type | 1.26 | 1.12 | 1.09 |
|  | 0.94, 1.69 | 0.83, 1.50 | 0.81, 1.47 |
|  | 0.125 | 0.459 | 0.566 |
| Evening type | 1.92 | 1.47 | 1.43 |
|  | 1.33, 2.76 | 1.01, 2.13 | 0.97, 2.10 |
|  | <0.001 | 0.043 | 0.072 |
| Sex |  |  |  |
| Men (Reference) | 1.00 | 1.00 | 1.00 |
|  | 1.00,1.00 | 1.00,1.00 | 1.00,1.00 |
|  | . | . | . |
| Women | 0.26 | 0.71 | 0.63 |
|  | 0.20, 0.34 | 0.52, 0.96 | 0.45, 0.87 |
|  | <0.001 | 0.027 | 0.005 |
| Alcohol consumption (grams/day) |  |  |  |
| Abstain |  | 0.16 | 0.17 |
|  |  | 0.07, 0.36 | 0.07, 0.41 |
|  |  | <0.001 | <0.001 |
| 1-10 (Reference) |  | 1 | 1 |
|  |  | 1.00, 1.00 | 1.00, 1.00 |
|  |  | . | . |
| 11-20 |  | 4.52 | 4.43 |
|  |  | 3.26, 6.28 | 3.17, 6.20 |
|  |  | <0.001 | <0.001 |
| 21-30 |  | 6.32 | 6.43 |
|  |  | 4.35, 9.19 | 4.40, 9.39 |
|  |  | <0.001 | <0.001 |
| 31-40 |  | 7.73 | 7.58 |
|  |  | 5.08, 11.76 | 4.91, 11.70 |
|  |  | <0.001 | <0.001 |
| 41 or more |  | 8.44 | 8.3 |
|  |  | 5.86, 12.14 | 5.72, 12.05 |
|  |  | <0.001 | <0.001 |
| Time at risk (person-years) | 551,124 | 551,124 | 551,124 |
| N of deaths | 312 | 312 | 312 |
| N of persons | 22,976 | 22,976 | 22,976 |

<sup>1</sup>ICD-10 codes F10, G312, G4051, G621, G721, I426, K292, K70, K860, K852, O354, P043, Q860, and X45; includes also corresponding ICD-9 and ICD-8 codes {Official Statistics of Finland: Causes of Death (e-publication). Helsinki, Statistics Finland [referred: 22.4.2021]}.

**Table S5.** Mortality from malignant neoplasm of larynx, trachea, bronchus and lung^1^: Hazard ratios, 95% confidence intervals and p-values from multivariable models for three models by chronotype and smoking status in 1981-2018.

|  | Sex-age adjusted | Sex, age and smoking adjusted | Adjusted for all covariates, but showing only smoking |
| --- | --- | --- | --- |
|  | HR<br>95% CI<br>p-value | HR<br>95% CI<br>p-value | HR<br>95% CI<br>p-value |
| Morning type (Reference) | 1 | 1 | 1 |
|  | 1.00, 1.00 | 1.00, 1.00 | 1.00, 1.00 |
|  | . | . | . |
| Some morning type | 1.06 | 1.07 | 1.09 |
|  | 0.82, 1.37 | 0.83, 1.38 | 0.84, 1.41 |
|  | 0.639 | 0.611 | 0.500 |
| Some evening type | 1.21 | 1.06 | 1.07 |
|  | 0.94, 1.56 | 0.82, 1.36 | 0.83, 1.37 |
|  | 0.132 | 0.662 | 0.618 |
| Evening type | 1.78 | 1.28 | 1.25 |
|  | 1.29, 2.46 | 0.93, 1.77 | 0.90, 1.74 |
|  | <0.001 | 0.130 | 0.182 |
| Sex |  |  |  |
| Men (Reference) | 1.00 | 1.00 | 1.00 |
|  | 1.00, 1.00 | 1.00, 1.00 | 1.00, 1.00 |
|  | . | . | . |
| Women | 0.26 | 0.73 | 0.71 |
|  | 0.20, 0.33 | 0.56, 0.94 | 0.54, 0.93 |
|  | <0.001 | 0.016 | 0.013 |
| Smoking status (Cigarettes Per Day) |  |  |  |
| Never (Reference) |  | 1 | 1 |
|  |  | 1.00, 1.00 | 1.00, 1.00 |
|  |  | . | . |
| Occasional |  | 1.24 | 1.08 |
|  |  | 0.29, 5.21 | 0.25, 4.61 |
|  |  | 0.771 | 0.915 |
| Former |  | 4.43 | 3.92 |
|  |  | 2.77, 7.09 | 2.46, 6.26 |
|  |  | <0.001 | <0.001 |
| Light (1-9 CPD) |  | 10.12 | 7.85 |
|  |  | 6.02, 17.01 | 4.68, 13.16 |
|  |  | <0.001 | <0.001 |
| Medium (10-19 CPD) |  | 23.24 | 17.57 |
|  |  | 15.21, 35.49 | 11.46, 26.94 |
|  |  | <0.001 | <0.001 |
| Heavy (20 or more CPD) |  | 35.94 | 27.5 |
|  |  | 23.09, 55.96 | 17.51, 43.18 |
|  |  | <0.001 | <0.001 |
| Time at risk (person-years) | 551,124 | 551,124 | 551,124 |
| N of deaths | 405 | 405 | 405 |
| N of persons | 22,976 | 22,976 | 22,976 |

<sup>1</sup>ICD-10 codes C32-C34, includes also corresponding ICD-9 and ICD-8 codes {Official Statistics of Finland: Causes of Death (e-publication). Helsinki, Statistics Finland [referred: 22.4.2021]}.

## Notes

Funding: JK is supported by the Academy of Finland (grant # 336823) and the Sigrid Juselius Foundation.

Conflicts of interest: None

### Competing Interest Statement

The authors have declared no competing interest.

### Funding Statement

the Academy of Finland (grant # 336823) and the Sigrid Juselius Foundation

